# NeuroBooster Array: A Genome-Wide Genotyping Platform to Study Neurological Disorders Across Diverse Populations

**DOI:** 10.1101/2023.11.06.23298176

**Authors:** Sara Bandres-Ciga, Faraz Faghri, Elisa Majounie, Mathew J Koretsky, Jeffrey Kim, Kristin S Levine, Hampton Leonard, Mary B Makarious, Hirotaka Iwaki, Peter Wild Crea, Dena G Hernandez, Sampath Arepalli, Kimberley Billingsley, Katja Lohmann, Christine Klein, Steven J Lubbe, Edwin Jabbari, Paula Saffie-Awad, Derek Narendra, Armando Reyes-Palomares, John P Quinn, Claudia Schulte, Huw R Morris, Bryan J. Traynor, Sonja W. Scholz, Henry Houlden, John Hardy, Sonya Dumanis, Ekemini Riley, Cornelis Blauwendraat, Andrew Singleton, Mike Nalls, Janina Jeff, Dan Vitale Global Parkinson’s Genetics Program and the Center for Alzheimer’s Disease and Related Dementias

## Abstract

Genome-wide genotyping platforms have the capacity to capture genetic variation across different populations, but there have been disparities in the representation of population-dependent genetic diversity. The motivation for pursuing this endeavor was to create a comprehensive genome-wide array capable of encompassing a wide range of neuro-specific content for the Global Parkinson’s Genetics Program (GP2) and the Center for Alzheimer’s and Related Dementias (CARD). CARD aims to increase diversity in genetic studies, using this array as a tool to foster inclusivity. GP2 is the first supported resource project of the Aligning Science Across Parkinson’s (ASAP) initiative that aims to support a collaborative global effort aimed at significantly accelerating the discovery of genetic factors contributing to Parkinson’s disease and atypical parkinsonism by generating genome-wide data for over 200,000 individuals in a multi-ancestry context. Here, we present the Illumina NeuroBooster array (NBA), a novel, high-throughput and cost-effective custom-designed content platform to screen for genetic variation in neurological disorders across diverse populations. The NBA contains a backbone of 1,914,934 variants (Infinium Global Diversity Array) complemented with custom content of 95,273 variants implicated in over 70 neurological conditions or traits with potential neurological complications. Furthermore, the platform includes over 10,000 tagging variants to facilitate imputation and analyses of neurodegenerative disease-related GWAS loci across diverse populations. The NBA can identify low frequency variants and accurately impute over 15 million common variants from the latest release of the TOPMed Imputation Server as of August 2023 (reference of over 300 million variants and 90,000 participants). We envisage this valuable tool will standardize genetic studies in neurological disorders across different ancestral groups, allowing researchers to perform genetic research inclusively and at a global scale.

## Introduction

Historically, commercial genome-wide genotyping arrays have been designed by selecting tag SNPs (SNP in a region of the genome with high linkage disequilibrium that represents an haplotype) from European or Asian populations. Little attention has been paid to designing arrays that can globally cover genetic variation across populations. The impetus behind our endeavor was to develop a comprehensive genome-wide array capable of encompassing a broad spectrum of neuro-specific content. This array serves the objectives of two key initiatives: the Global Parkinson’s Genetics Program (GP2) and the Centre for Alzheimer’s and Related Dementias (CARD).

Much has been done to unravel the genetic landscape of brain disorders, identifying both causal and risk variants underlying disease risk and progression in a wide range of neurological diseases. Despite these recent efforts there is an unmet need to address: the vast majority of genetic studies have focused on populations of European ancestry ^1,2^. The genetic architecture of other populations, such as Africans, Asians and Latino, remains largely unexplored in neurology and other areas of research with only few studies being published ^3,4,5,6,7,8^.

Here, we present the NeuroBooster Array v.1.0 (NBA), a cost-efficient platform to perform ancestry-inclusive Genome-Wide Association Study (GWAS) and rare variant detection. The NBA represents a valuable screening tool to rapidly identify risk variants and disease modifiers that contribute to neurological conditions across ancestries, as well as known causal pathogenic variants with almost eight times the coverage of previous platforms like Neurochip ^9^ and NeuroX arrays ^10^. The NBA is a reliable platform to accelerate participant enrollment for target-specific clinical trials according to underlying genetics, as well as sample prioritization for more costly approaches such as whole-genome sequencing and long-read sequencing.

In the present manuscript, we describe the NBA custom design, due diligence, and content. Furthermore, we discuss coverage and imputation accuracy for common and rare variation in the context of previous arrays (NeuroX ^10^ and NeuroChip ^9^). We comprehensively evaluate the NBA’s performance by assessing SNP imputation via mean imputed r^2^ and minor allele frequency across 1000 Genomes populations data, including Americans (AMR), African Americans (AAC), Africans (AFR), East Asians (EAS), South Asians (SAS) and Europeans (EUR), using several population reference panels such as the TOPMed Imputation Server (https://imputation.biodatacatalyst.nhlbi.nih.gov), the Haplotype Reference Consortium r1.1 2016 (HRC) (http://www.haplotype-reference-consortium.org), the Genome Asia Pilot Project (GASP) (https://genomeasia100k.org/) and the Consortium on Asthma among African-ancestry Populations in the Americas (CAAPA) Panel (https://www.caapa-project.org/). Finally, we discuss the benefits and limitations of using this platform to study polygenic inheritance in neurological conditions.

## Methods

### NeuroBooster array design

The NBA contains a backbone of 1,914,934 genetic markers from the Infinium Global Diversity Array-8 v1.0 (GDA) complemented with custom content of 95,273 disease-associated variants involved in a wide range of neurological conditions. The custom content on the NBA was purposely designed to complement the base content of the GDA series of arrays from Illumina **(Table 1).**

**Table 1.** Coverage differences between NeuroX, NeuroChip and NeuroBooster arrays.

|  |  |  |  |  |  |
| --- | --- | --- | --- | --- | --- |
| Variant description | NeuroBooster | NeuroChip | Comparison with Neurochip (+) | NeuroX | Comparison with NeuroX (+) |
| Total variants (pre-QC) | 2.010.207 | 486.137 | 1.524.070 | 267.607 | 1.742.600 |
| Backbone | 1.914.934 | 306.670 | 1.608.264 | 242.901 | 1.672.033 |
| Custom-content variants | 95.273 | 179.467 | -84.194 | 24.706 | 70.567 |
| Indels | 49.554 | 16.259 | 33.295 | 200 | 49.354 |
| Autosomal variants | 1.873.290 | 473.442 | 1.399.848 | 261.477 | 1.611.813 |
| Coding variants | 1.223.002 | 88.560 | 1.134.442 | 226.104 | 996.898 |
| Sex chromosomal variants | 79.994 | 11.840 | 68.154 | 5.906 | 74.088 |
| Mitochondrial variants | 1.509 | 160 | 1.349 | 219 | 1.290 |
| Variants with MAF < 0.05* | 853.885 | 227.448 | 626.437 | 219.093 | 634.792 |
| Variants with MAF < 0.001* | 148.821 | 154.953 | -6.132 | 179.500 | -30.679 |
| * Based on annotations |  |  |  |  |  |

The GDA is built on a high-density SNP global backbone optimized for cross-population imputation coverage of the genome [https://www.illumina.com/products]. The GDA contains a robust genome-wide scaffold designed to tag both common and low frequency variants (minor allele frequency > 1%) in global populations. This scaffold was designed through collaborations with the Consortium on Asthma among African-ancestry Populations in the Americas (CAAPA) [https://www.caapa-project.org/] and the Population Architecture using Genomics and Epidemiology (PAGE) study [https://www.pagestudy.org/]. For GDA design, more than 400,000 markers of exome content were gathered from 36,000 individuals of diverse ethnic groups, including African Americans, Hispanics, Pacific Islanders, East Asians, and individuals of mixed ancestry. The GDA includes content not found in the 1K Genomes Project, with more than 1000 whole-genome sequences of African ancestry and populations throughout the Americas, including the US, Caribbean, and Latin and South America. The GDA is designed to contain diverse exonic content from, Genome Aggregation Database version 2.1 (https://gnomad.broadinstitute.org/news/2018-10-gnomad-v2-1/, RRID:SCR_014964) including both cross-population and population-specific markers with either functionality or strong evidence for association with certain populations (*Illumina Inc)*.

We designed the custom content for the NBA with four goals in mind. These goals included: 1) identify rare coding variants associated with disease of interest to researchers in the neurology field; 2) improve imputation of known GWAS loci for neurodegenerative diseases across populations of diverse ancestries; 3) generally improve imputation quality across populations of diverse ancestries to facilitate risk locus discovery; and 4) facilitate fine-mapping of risk loci.

To accomplish these aims, we aggregated custom content from a variety of neurodegenerative disease-relevant sources. In brief, building the array was carried out in five phases (described further below), consisting of sequential design components. The first design component was a systematic review as well as consultation with key opinion leaders in the field of neurology to identify putative rare variants associated with disease. The second design component consisted of annotating known GWAS risk loci and identifying multi-population tag SNPs to support improved imputation and locus tagging across ancestry groups in follow-up studies. Based on this information, the third component consisted of saturation of genotyping of risk loci from large GWAS regions. Next, we carried out probe design due diligence in collaboration with *Illumina Inc*. Finally, we used diverse samples and rare-variant-enriched cohorts to build a custom cluster file to improve genotype quality. The culmination of these phases of work allowed us to densely genotype and impute higher variant coverage into regions of interest related to neurological diseases.

### Systematic review of publicly annotated databases and key opinion leaders

Whole-exome and whole-genome sequencing variant prioritization analyses were conducted in families and cohort studies. This includes some from diverse ancestries and population isolates, provided by International Parkinson’s Disease Genomics Consortium (IPDGC, https://pdgenetics.org) members and collaborators and used as the first layer of derived content nominated by key opinion leaders. We followed this up with systematic reviews of publicly annotated databases, including the Human Gene Mutation Database (HGMD) version 2019 (http://www.hgmd.org/, RRID:SCR_001621) ^11^ and the Genomics England panel (accessed in May 2019) (https://panelapp.genomicsengland.co.uk/).

From HGMD, we extracted any nucleotide substitutions (missense/nonsense, regulatory, splicing) and also variants labeled as nucleotide deletions, insertions and indels for any disease or phenotype combinations suggested by key opinion leaders as potentially being implicated in the etiology of neurological diseases or conditions presenting with neurological complications. Our exhaustive list of search terms are highlighted in **Supplementary Table 1**. The database cross-referencing resulted in nearly 90,000 candidate variants initially. Due to array design and space concerns on the product, the variant list from this first portion of the design was reduced to 54,907 variants prior to Illumina due diligence (described later, **Supplementary Table 5)**.

We also included an allocation of 1,809 polygenic risk score (PRS)-related variants for Parkinson’s disease risk nominated by PRSice-2 Version 2.2.13 ^12^ (https://choishingwan.github.io/PRSice/, DOI: 10.5281/zenodo.3703335, RRID:SCR_017057). This includes variants that did not reach genome-wide significance but positively impacted PRS modeling efforts in previous reports. For more details on PRS construction, please refer to Nalls et al. 2019.

### Identification of multi-ancestry tag SNPs from GWAS loci

Loci identified as genome-wide significant in large-scale GWAS meta-analyses of neurodegenerative diseases ^13, 14, 15, 4, 16, 17, 18, 19^ were the next target of our design process. Genome-wide significant loci from eight neurodegenerative disease-related conditions, including Alzheimer’s disease, Parkinson’s disease, amyotrophic lateral sclerosis, multiple sclerosis, progressive supranuclear palsy, corticobasal degeneration, multiple system atrophy, frontotemporal dementia and dementia with Lewy bodies were included with either multiple proxies for the top SNP at every locus, or technical replicates when proxies were not available. GWAS tagging SNPs per locus were defined as any SNP reaching a genome-wide significant p-value and correlated at r^2^ < 0.3 with any other significant SNPs within 250 kilobases. This process was done for each disease of interest based on the combined European reference series from 1000 Genomes phase 1v5 data (https://www.internationalgenome.org/data-portal/data-collection/phase-1,RRID:SCR_0068 28) after filtering all variants for “PASS” annotation and minor allele count of at least three. This led to the successful inclusion of ∼180 GWAS-related variants **(Supplementary Table 2)**.

To build multi-population references, the 1000 Genomes sequence data was again subsetted into ancestry reference “sub-populations”. These included conglomerate datasets from population labels as follows: MXL (Mexican Ancestry), CLM (Colombian), PEL (Peruvian), and PUR (Puerto Rican) were combined to form the AMR (American admixed) ancestry group; JPT (Japanese), CDX (Chinese Dai in Xishuangbanna), CHB (Han Chinese in Beijing), CHS (Han Chinese South), KHV (Kinh in Ho Chi Minh City, Vietnam) and CHD (Han Chinese in Beijing) were combined to form the EAS (East Asian) ancestry group; TSI (Toscani in Italy), IBS (Iberian Populations in Spain), GBR (British From England and Scotland), and CEU (Central European) were combined to form the EUR (European) ancestry group; PJL (Punjabi in Lahore, Pakistan), ITU (Indian Telugu in the U.K), STU (Sri Lankan Tamil in the UK), GIH (Gujarati Indians in Houston, Texas, USA), BEB (Bengali in Bangladesh) were combined to form the SAS (South Asians) ancestry group; GWD (Gambian in Western Division – Mandinka), MSL (Mende in Sierra Leone), ESN (Esan in Nigeria), YRI (Yoruba in Ibadan, Nigeria), LWK (Luhya in Webuye, Kenya), GWJ (Gambian Jola), GWF (Gambian Fula), and GWW (Gambian Wolof) were combined to form the AFR (African) ancestry group; ASW (African Ancestry in SW USA) and ACB (African Caribbean in Barbados) were combined to form the AAC (African admixed) ancestry group. Due to sample size issues for tagging analysis, a total of 115 samples from Finland were excluded from the references leaving us with 3,395 reference samples across all groups. Based on these reference samples and the 180 GWAS loci discussed earlier, the software TagIt version 1.0.8 https://bioinformaticshome.com/tools/imputation/descriptions/TagIt.html^20^ was run using default settings per locus stratified by ancestry group to identify diverse tag SNP series (r2 > 0.3).

### Saturation genotyping of known risk loci5

For each of the 180 GWAS variants of interest tagging unique loci, we identified upstream and downstream tag SNPs across all ancestry groups. For each region, all tag SNPs from diverse populations were included. To fill out content after the Illumina design triage described below in the due diligence content section, a surplus of EUR ancestry tag SNPs per locus were added. This phase of design included over 10,000 SNPs tagging diverse ancestral linkage structure at all loci. Regions tagged by the multi-population variants of interest covered over 38MB of the genome.

### NeuroBooster array due diligence content

Illumina design scoring was undertaken at *Illumina Inc* to screen potential array content. This included a scoring of variants for quality and their ability to be included on arrays. Variants whose probes were problematic and for which low scoring could not reliably be built were excluded from the beadpool. Additionally, after clustering 2,793 diverse samples from the Global Parkinson’s Genetics Program ^21^ [https://gp2.org/training/] across multiple populations **(Supplementary Table 3)**, we reviewed variants that were not accurately genotyped, with high rates of missingness across batches resulting in the exclusion of 49,590 variants **(Supplementary Table 4).** After iterative design scoring at *Illumina Inc*, custom content for 95,273 variants (and the additional 1,914,934 standard content variants) were annotated using the Variant Effect Predictor Release 100 and made available as part of the standard series of GP2 releases [https://useast.ensembl.org/info/docs/tools/vep/index.html].

### NeuroBooster array data processing and custom clustering

Automated genotype data processing was conducted on GenoTools, a Python pipeline built for quality control and ancestry estimation of data. Additional details can be found at (https://github.com/GP2code/GenoTools). Quality control (QC) was performed according to standard protocols. Samples with a call rate below 95%, sex mismatches, or high heterozygosity (estimated by an |F| statistics of > 0.25) were excluded from analyses.

Further QC measures included the removal of SNPs with missingness above 5%, variants with significant deviations from Hardy-Weinberg Equilibrium (HWE P value < 1E-4), variants with non-random missingness and variants with missing data patterns (haplotype at P ≤ 1E-4 per ancestry). A total of 2,793 diverse case samples across six ancestry groups were clustered using the Illumina’s GenomeStudio Software v2.0.5 package (https://support.illumina.com/downloads/genomestudio-2-0.html, RRID:SCR_010973) **(Figure 1, Supplementary Table 3)**.

**Figure 1.**
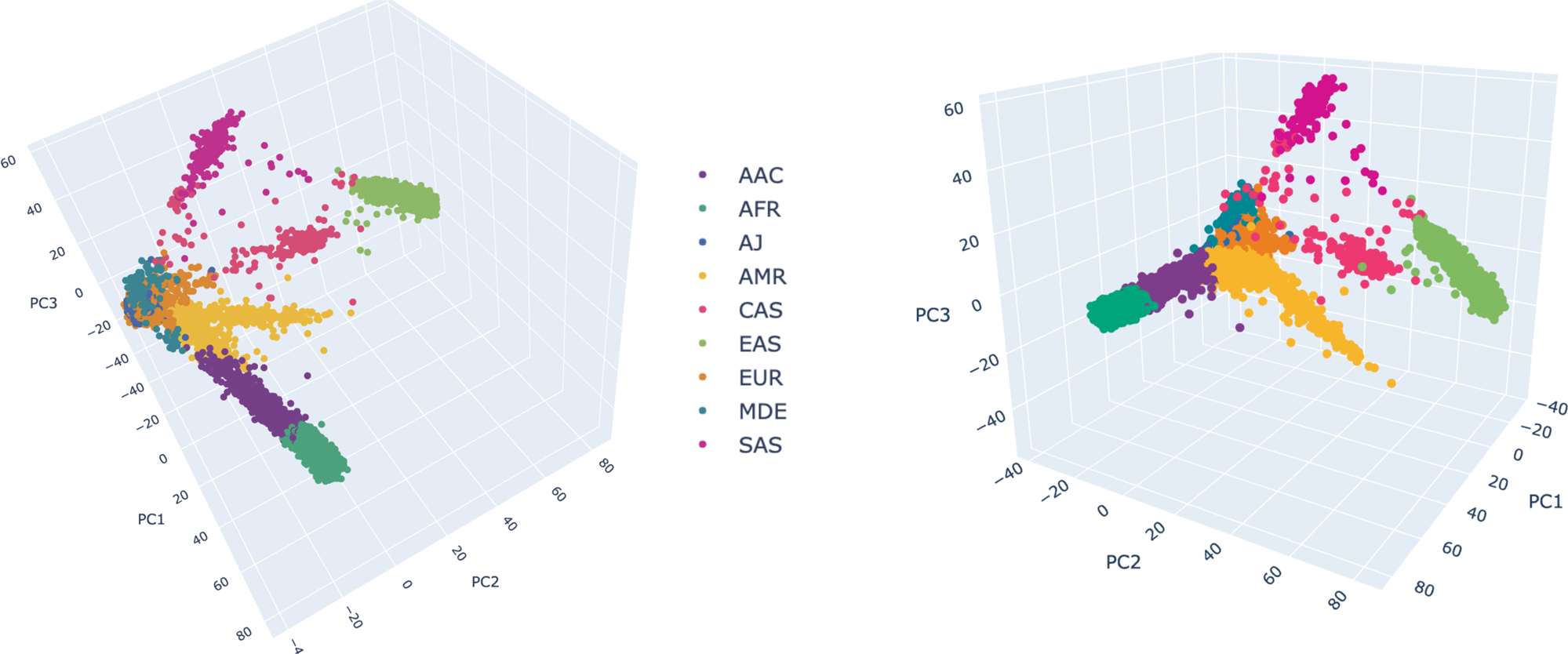
Ancestry prediction clustering for samples genotyped on the NeuroBooster array. 3 dimensional principal components analysis to group individuals based on their genetic makeup. A total of 2,793 samples from the Global Parkinson’s Genetics Program were included, including 2,373 Parkinson’s disease cases and 420 Gaucher disease cases. Each point represents a sample and the colors depict the ancestral background as shown in the color legend: EUR = orange, EAS = lemon green, AMR = yellow, AJ = lapis blue, AFR = teal blue, AAC = purple, SAS = magenta, CAS = dark pink, MDE = cerulean blue. EUR (Europe), EAS(East Asian), AMR (Latino/Admixed American), AJ (Ashkenazi Jewish), AAC (African-Admixed), AFR (African), SAS (South-Asian), CAS (Central-Asian), MDE (Middle East).

**Figure 2.**
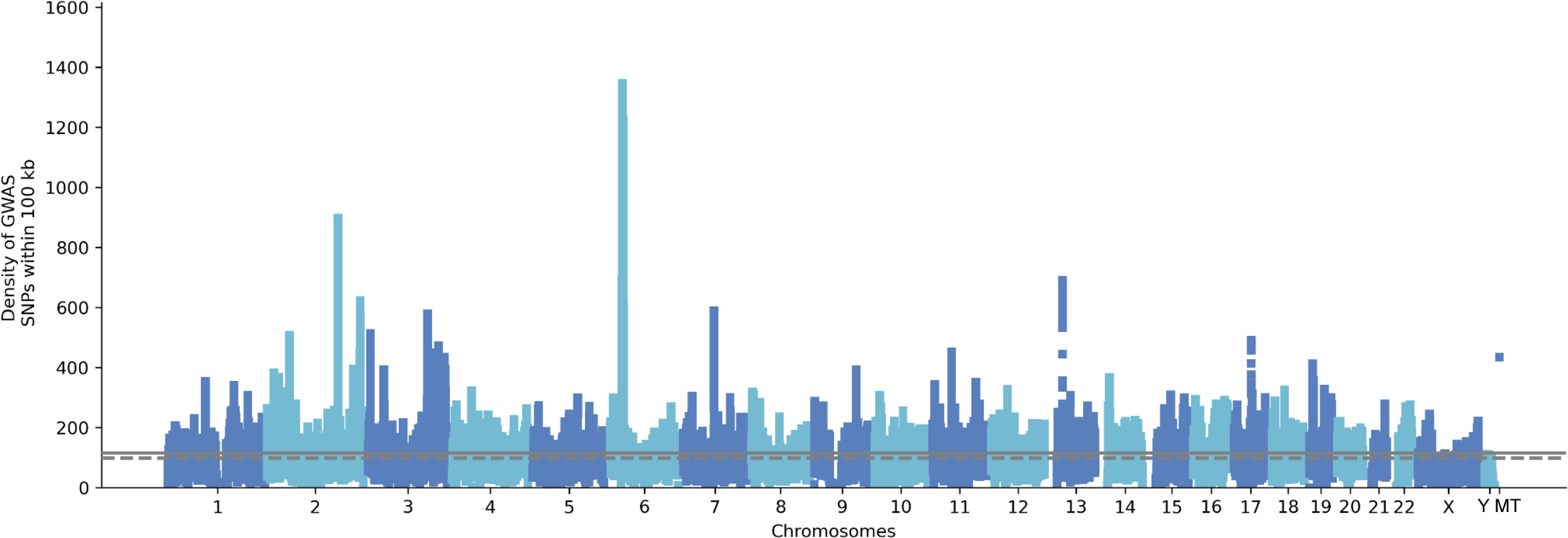
Brisbane plot showing the genomic density of SNPs on GP2 raw genotyped data generated on the NeuroBooster array.

The clustering file is available for download via the GP2 Github repository (https://github.com/GP2code; DOI 10.5281/zenodo.7904832, release 5; https://gp2.org/). Clusters can be viewed using the GP2 data browser. Among the 2,793 samples, a total of 2,373 Parkinson’s disease patients were included in addition to 420 Gaucher disease cases to capture variants of interest in the *GBA1* gene.

Clustering was done following *Illumina Inc* guidelines. Specifically, the robustness and reliability of genotype clustering was ensured by implementing a meticulous quality control protocol on the genotyping data. The protocol started with an evaluation of BeadChip performance in GenomeStudio using the Controls Dashboard. Any Beadchips with profiles suggestive of technical issues were promptly excluded from the analysis to negate the influence of technical inaccuracies on the results. Next, an assessment of sample call rates was conducted utilizing the Samples Table. To uphold the standard of genotyping data, samples presenting call rates lower than 0.98 were excluded from the subsequent analyses.

Following the assessment of call rates, an in-depth review of the Copy Number (CN) Metrics report was carried out to detect samples with outlier values for ‘LogRDev’ or ‘BAlleleDev’. Such samples, suggestive of potential genotyping anomalies, were excluded from further investigation to ensure a dataset free from major genotyping errors and anomalies. By employing this stringent sample inclusion/exclusion protocol, we assured that the subsequent genotype clustering was based on high-quality samples free from technical errors. This approach ensures that the genotyping data used for clustering is accurate, reliable, and most importantly, trustworthy.

Passing samples were then reclustered using the default *Illumina Inc* cluster algorithm built into GenomeStudio, with autosomes clustered together and sex chromosomes done separately.

### NeuroBooster array genotyping protocol

An overview of the protocol can be visualized in **Supplementary Figure 1**. Briefly, for each sample a total of 200 ng of high-quality genomic DNA is amplified and enzymatically fragmented. The resulting fragments are alcohol-precipitated and resuspended in a buffer. Next, the fragmented DNA solution is hybridized to the NBA using a Tecan Freedom EVO liquid-handling robot (Tecan, Research Triangle Park, NC). After hybridization, automated allele-specific, enzymatic base extension and fluorophore staining are performed. The stained genotyping arrays are washed, sealed and vacuum-dried prior to scanning them on the Illumina iScan system. Raw data files are imported into GenomeStudio (version 2.0, Genotyping Module, Illumina Inc) using a custom-generated sample sheet, and genotypes are called using a GenCall threshold of 0.15.

## Results

### NeuroBooster content overview

In total, the NBA contains 1,873,290 autosomal variants, 79,994 sex chromosomal variants (X and Y chromosomes), and 1,509 mitochondrial variants **(Figure 2)**. The overlap between the NBA and NeuroChip is n= 126,220 variants. The NBA includes over 10,000 multi-ancestry GWAS locus tagging variants to facilitate imputation and analyses of these neurodegenerative disease-related GWAS loci across diverse populations. Detailed pathogenic inferences and annotated NBA functionally variant content for the array in total is provided through GP2 ^21^. **Figure 3**, **Figure 4, and Supplementary Table 5** display the abundance of disease-associated variants per gene covered across the most prevalent neurodegenerative diseases based on initial systematic review and cross-referencing of the HGMD and Genomics England database content.

**Figure 3.**
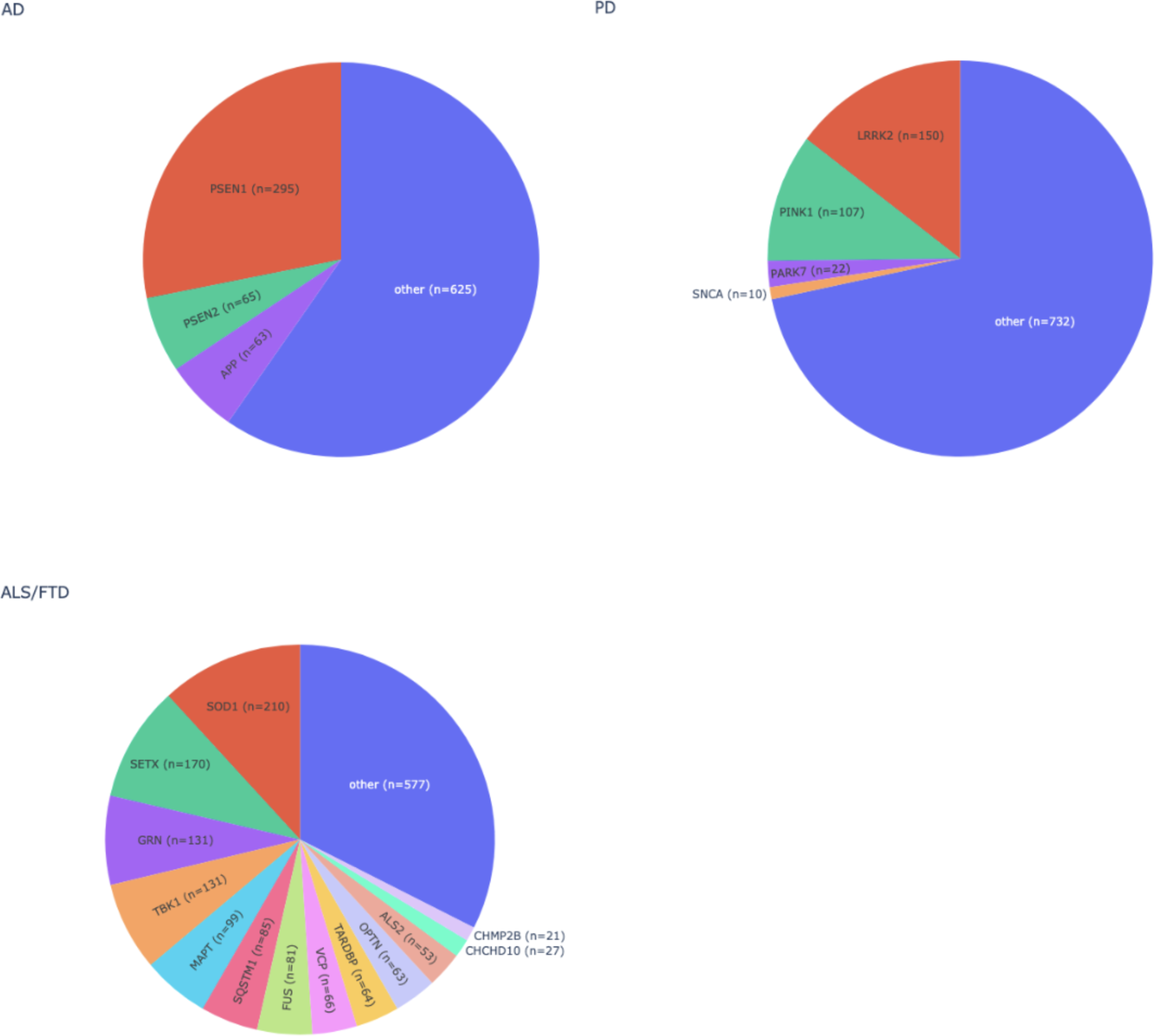
Overview of the number of Human Gene Mutation Database disease associated variants that are present on the NeuroBooster array for the most prevalent neurodegenerative diseases. AD = Alzheimer’s disease, ALS = amyotrophic lateral sclerosis, FTD = frontotemporal dementia, and PD = Parkinson’s disease

**Figure 4A.**
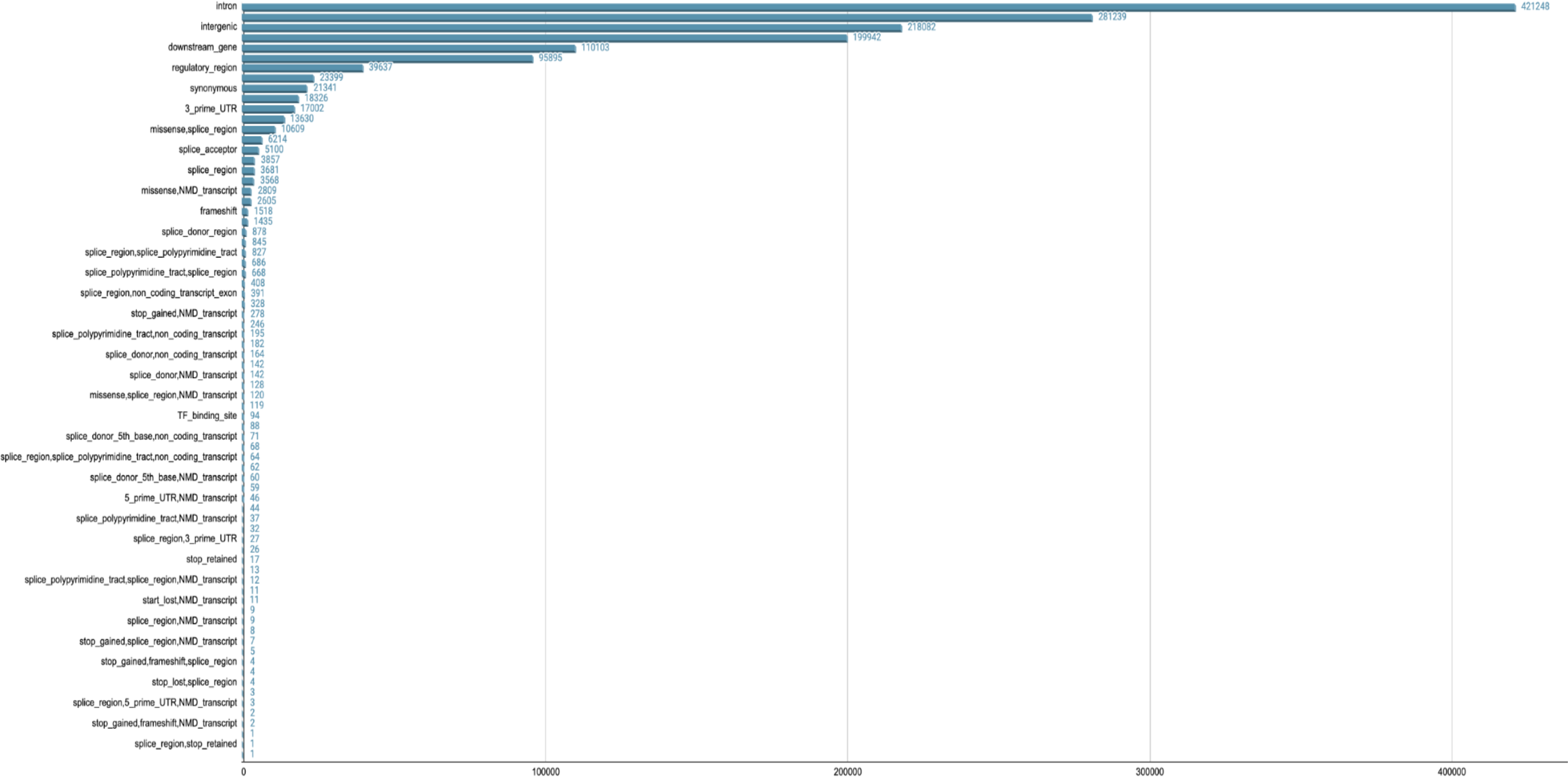
Overview of NeuroBooster array content by variant category

**Figure 4B.**
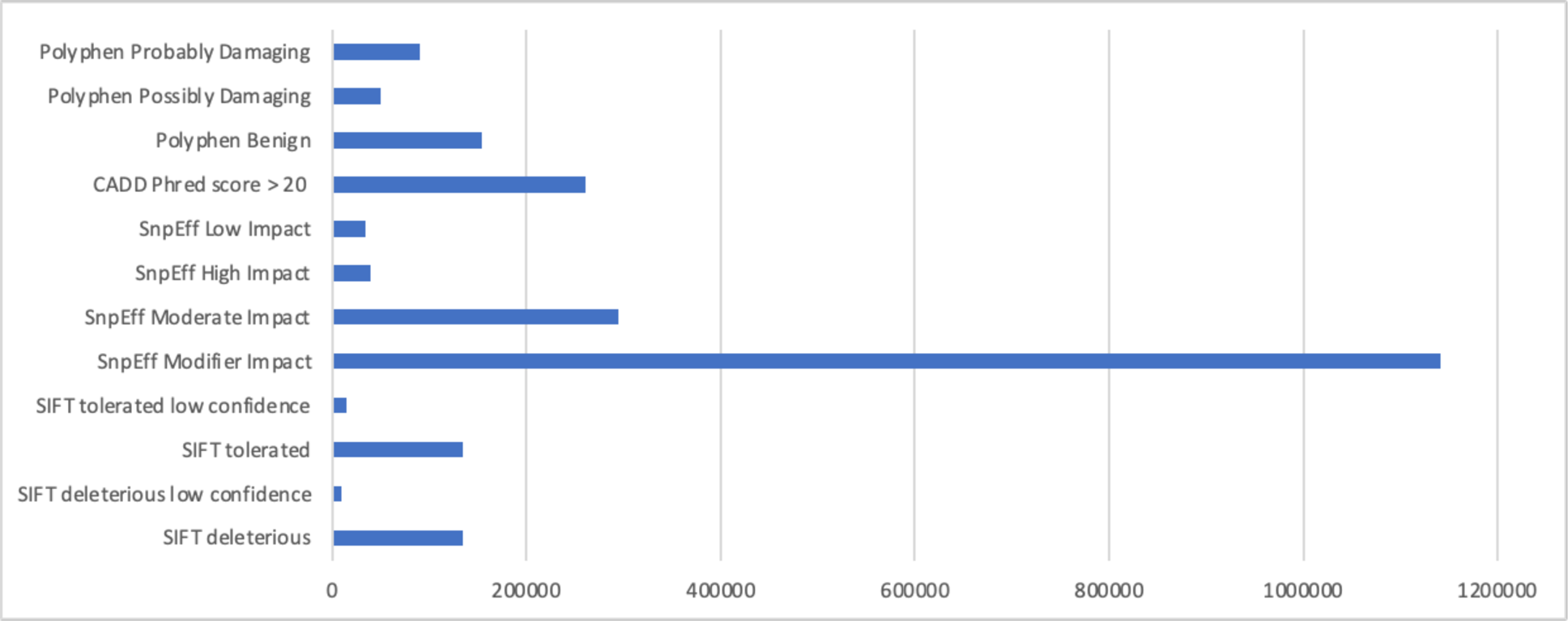
Overview of NeuroBooster array content by pathogenicity predictors

### Genotyping accuracy

GenTrain scores were calculated for all NeuroBooster variants using GenomeStudio version 2.0. The GenTrain score is a statistical score based on the shapes of the different allelic clusters and their relative distance to each other (Illumina). Typically, GenTrain scores above 0.7 are considered high quality genotypes. The mean (and SD) for the array is 0.83 (0.091) **(Supplementary Figure 2**). Due to improved technology from *Illumina Inc* compared to NeuroChip or NeuroX arrays over the past decade as well as the iterative design process the array underwent in collaboration with *Illumina Inc*, we are able to rescue many variants not previously able to be genotyped (see **Supplementary Figure 3** for example comparison).

Genetic variants in *APOE, APP, GBA1, LRRK2, PARK2, DJ-1, PINK1, PRKN, PSEN1, SNCA, TREM2* and *VPS35* contribute to AD, LBD and PD etiology among other neurodegenerative diseases. Genotyping of some of these genes remains complicated due to high GC content (*APOE*) or the presence of a pseudogene (*GBA1*). We comprehensively assessed variant content for tagging and imputed variants across multiple ancestries (additional details can be found in **Supplementary Table 6, Supplementary Table 8).**

### Quality control metrics for imputation accuracy and allele frequencies intervals across diverse populations

Variants with a minor allele frequency (MAF) < 0.005 and Hardy-Weinberg equilibrium (HWE) *p*-value < 1E-5 were excluded before submission to the TOPMed Imputation server. The utilized TOPMed reference panel version, known as r2, encompasses genetic information from 97,256 reference samples and over 300 million genetic variants across the 22 autosomes and the X chromosome. As of October 2022, the TOPMed panel includes approximately 180,000 participants, with 29% of African, 19% of Latino/Admixed American ancestry, 8% of Asian ancestry, and 40% of European ancestry (https://topmed.nhlbi.nih.gov/). Further details about the TOPMed Study, Imputation Server, and Minimac Imputation can be accessed at https://topmed.nhlbi.nih.gov/, https://imputation.biodatacatalyst.nhlbi.nih.gov/#!(RRID:SCR_009292), https://genome.sph.umich.edu/w/index.php?title=Minimac:_Tutorial&oldid=14562, (RRID:SCR_009292). Following imputation, the resulting files underwent pruning based on a minor allele count (MAC) threshold of 10 and an imputation Rsq value of 0.3.

Imputation of autosomal variants was performed across 2,793 individuals across ten different ancestries **(Supplementary Table 7).** In its application to recent GP2 dataset releases [https://gp2.org/], imputation yields were solid. After filtering for imputation quality (r2 > 0.3 and minor allele count > 10) across ancestry groups, minimum variant yields using TOPMed imputation included 7.4 M to 19.6 M variants in total across ancestry groups. Lower frequency variants at MAF < 5% were also well imputed in this new data resource, with yields after filtering ranging from 0.8 M to 7.1 M across populations.

In addition to data imputation using TOPMed, we next assessed imputation accuracy performance by extracting the variant content available on the NBA from whole genome sequencing data from six ancestral populations available on 1000 Genomes (AAC, AFR, AMR, EAS, EUR, SAS). Optimal array design depends not only on tag SNP selection but also on empirical evaluation of imputation performance ^20^. We compared imputation performance for neurodegenerative diseases related GWAS loci with and without the custom NBA content. For each of the population-specific GWAS scaffolds, imputation accuracy was assessed by MAF bins versus various imputation reference panels, including Haplotype Reference Consortium (HRC), the Consortium on Asthma among African-Ancestry Populations in the Americas (CAAPA) the Genome Asia Pilot (GAsP) **(Supplementary Figure 4A-H)**.

## Discussion

Genetics research needs to expand from European to non-European population. By filling this critical disparity, we will build a holistic picture of neurological diseases in the global population. The NBA represents a cost-effective and powerful tool for researchers to test and validate genetic associations in a multi-ancestry context, delivering insights for both discovery and screening applications. This effort demonstrates the power of modern genomic technologies to rapidly screen large sample collections and to unravel the genetic architecture underlying diverse neurodegenerative diseases in a multi-ancestry context.

The availability of data from diverse populations generated through NBA can directly lead not only to the identification and replication of identified GWAS loci, but also help to narrow the search window within loci that had been previously discovered in the European population. The genome-wide assessment of human populations is useful in providing insights on the evolutionary history of the human genome, with a special emphasis on refining genetic associations and disease fine-mapping. Given the genetic diversity that exists across populations, an accurate screening represents a valuable resource to provide insights into genetic determinants underlying disease risk and progression, modifiers of disease, potential differences in the heritable component of disease between groups, and the generation of population-specific genetic risk profiles. The goal is ultimately to drive transformational progress in our understanding of the genetic architecture of neurological diseases in a global context that can be of benefit to patients in all populations.

A crucial aspect is the use of NBA to generate predictive information that can play a significant role in improving the design of clinical trials. This may enable the support of trials even in preclinical participants, allowing mechanistic stratification of disease and adjustments in trial outcomes based on the personalized predictions of disease for each individual.

However, it is essential to recognize the limitations inherent in this technology. Like all genotyping arrays, NBA does not detect DNA variants that were unknown prior to its design in 2019 as well as ultra-rare variants. It also cannot genotype variants in complex genomic regions (such as those involving pseudogenes) or identify repeat expansions or small structural variants due to the challenges in designing reliable probes. Our custom content is based on known-disease related genes and we assume the limitation that novel genetic disease contributors may be missing in this first version of the array.

In conclusion, we describe the design and implementation of the NBA, which offers more comprehensive and improved content compared to its predecessor platforms. The NBA serves as an invaluable asset in our quest to comprehend neurological diseases within a worldwide framework, particularly as we embark on the era of precision therapeutics.

## Supporting information

Supplementary authors

## Data availability

Data (DOI 10.5281/zenodo.7904832, release 5) used in the preparation of this article were obtained from the Global Parkinson’s Genetics Program (GP2) and can be accessed at amp-pd.org.

## Code availability

https://github.com/GP2code/Neuro_Booster_Array (DOI 10.5281/zenodo.10018765)

---

**Supplementary Figure 1.** Overview of NeuroBooster Array genotyping protocol

**Supplementary Figure 2.** GenTrain scores of the NeuroBooster divided by minor allele frequency.

**Supplementary Figure 3.** Cluster plot comparison of NeuroChip versus NeuroBooster array probes for LRRK2 p.Thr1410Met.

**Supplementary Figure 4A-H.** Imputation accuracy of tag GWAS hits across diverse populations from 1000 genomes data using diverse imputation panels

**Supplementary Table 1.** Neurological conditions/traits with potential neurological complications

**Supplementary Table 2.** Top risk variants nominated by recent GWAS meta-analyses across neurodegenerative diseases: Imputation qualities across multiple ancestries, average and standard deviation of the number of variants per locus

**Supplementary Table 3.** Custom genotype clustering: Overview of genotyped and imputed samples divided by ancestry

**Supplementary Table 4.** Problematic or low quality variants excluded from the array design

**Supplementary Table 5.** Neurobooster annotated content through the Human Gene Mutation Database

**Supplementary Table 6.** Imputed genetic markers for genes linked to neurodegenerative diseases across multiple ancestries

**Supplementary Table 7.** Number of imputed markers by minor allele frequency across different ancestral populations

