## Supplementary Figures for "NeuroBooster Array: A Genome-Wide Genotyping Platform to Study Neurological Disorders Across Diverse Populations"

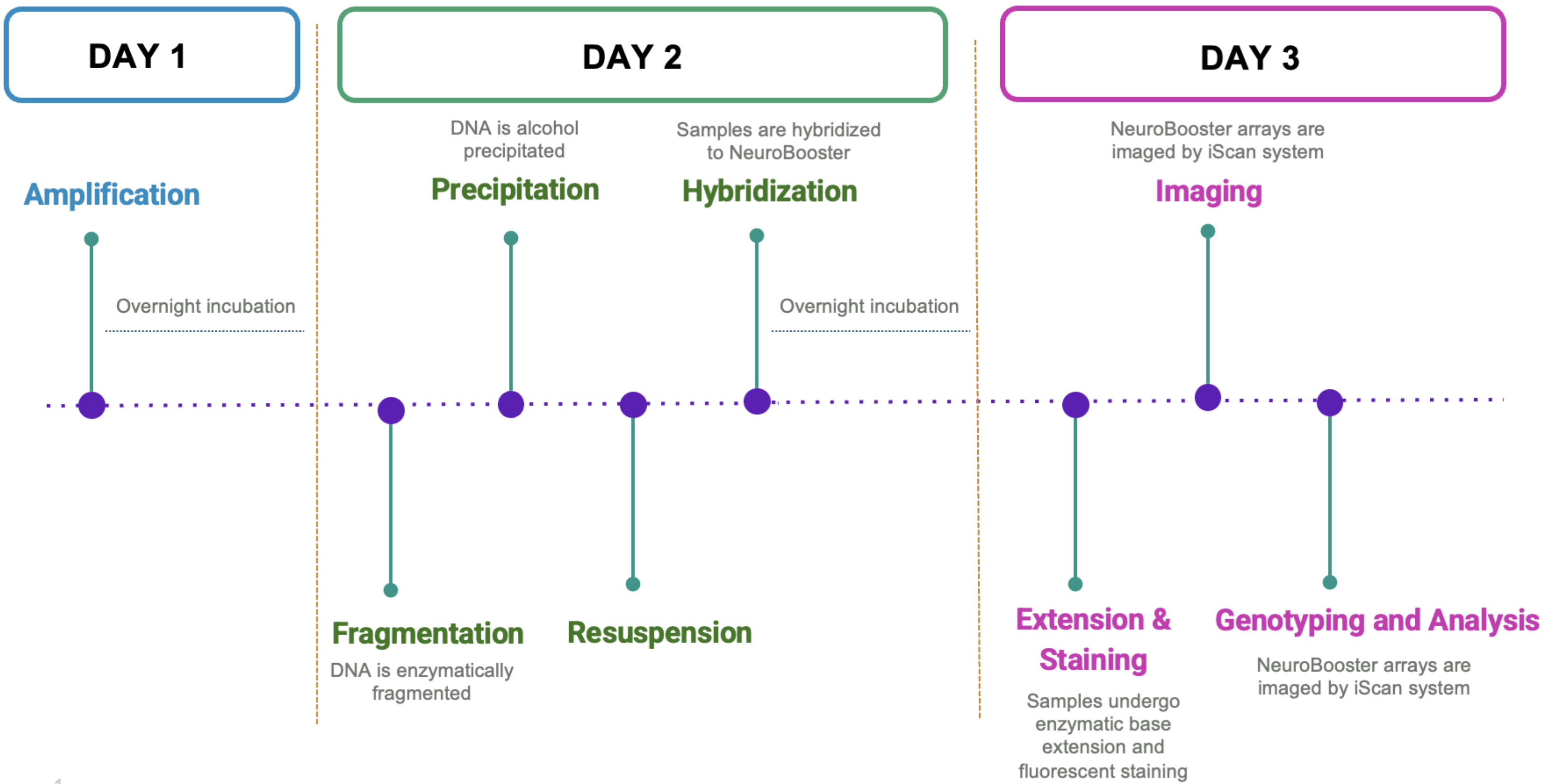


1

**Supplementary Figure 1. Overview of NeuroBooster Array genotyping protocol**

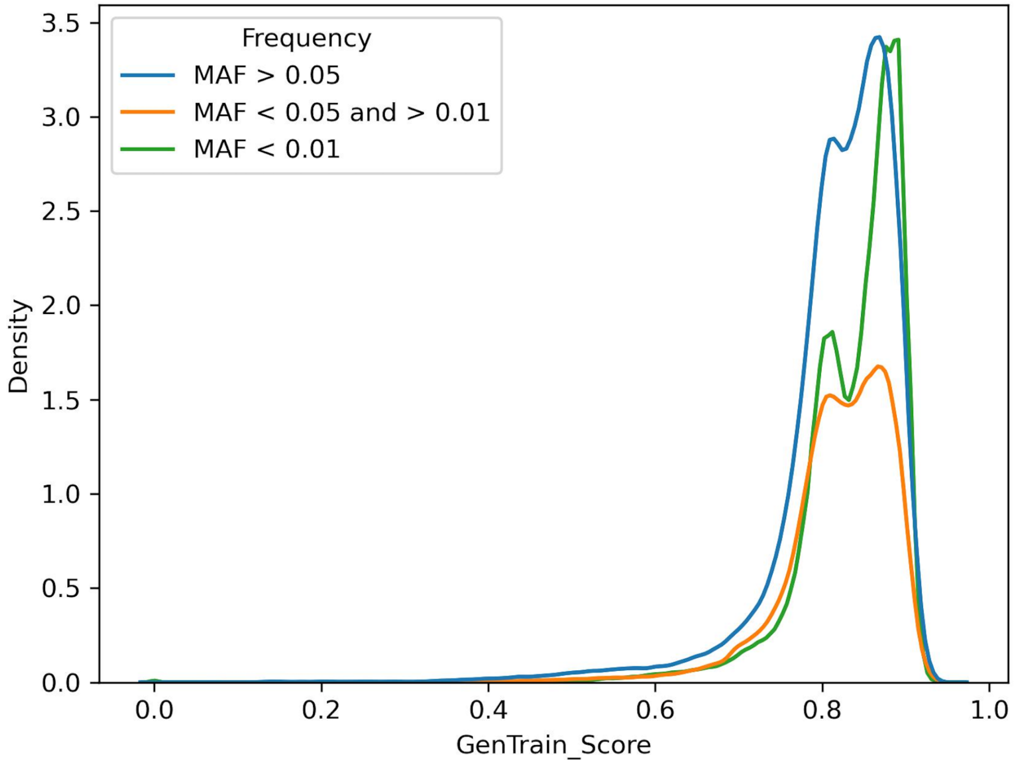


**Supplementary Figure 2. GenTrain scores of the NeuroBooster divided by minor allele frequency.** NeuroBooster variants were divided in three group by minor allele frequency (MAF): larger than 5%, between 5-1% and lower than 1%.

A)

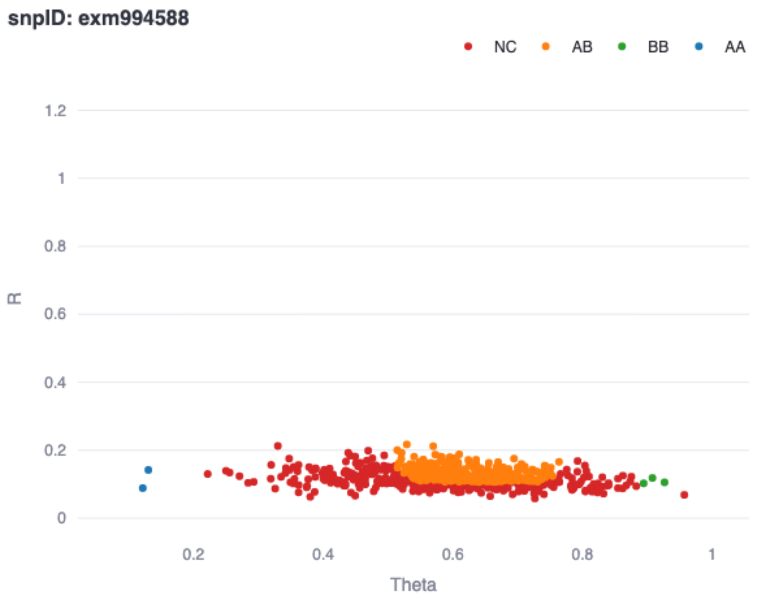


B)

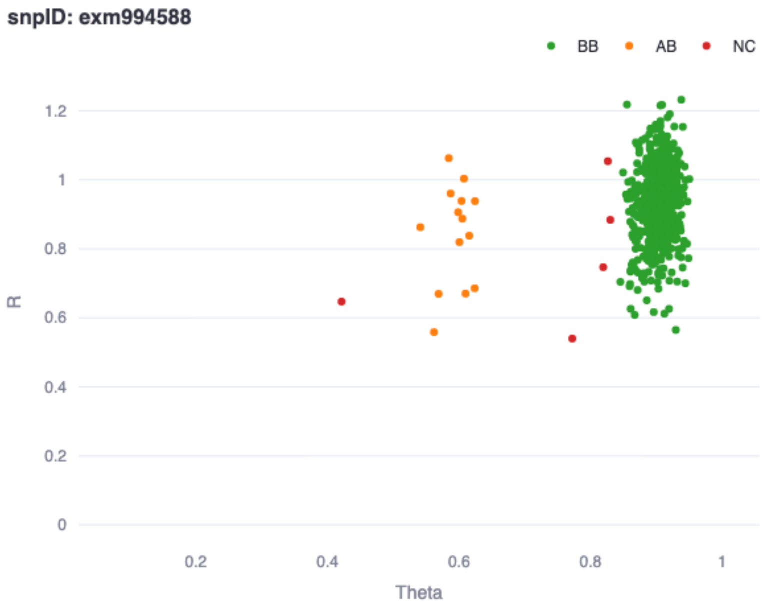


**Supplementary Figure 3. Cluster plot comparison of NeuroChip versus NeuroBooster array probes for *LRRK2* p.Thr1410Met.**
A) chr12:40309145 probe on NeuroChip array B) chr12:40309145 probe on NeuroBooster array

A)

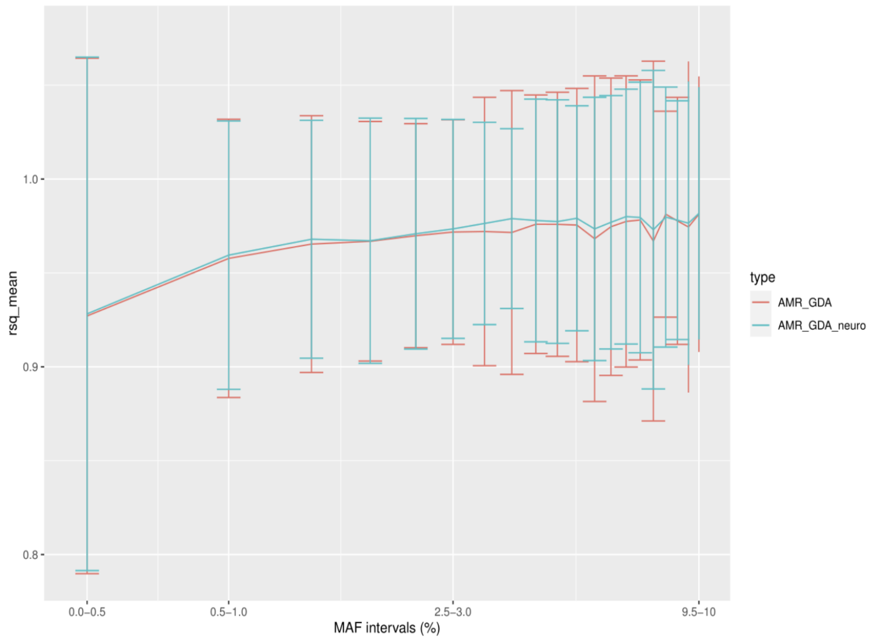


B)

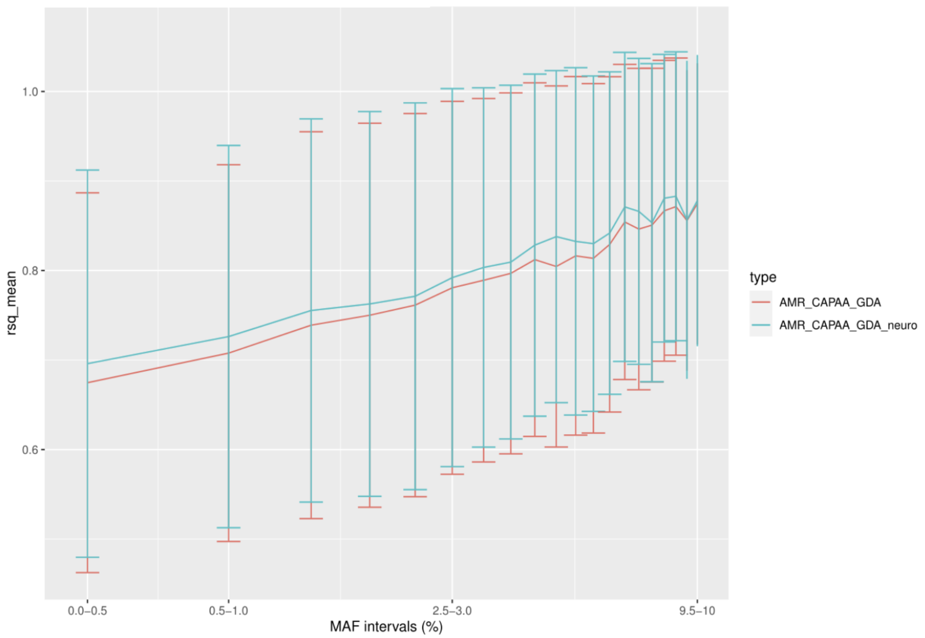


**Supplementary Figure 4A-H. Imputation accuracy of tag GWAS hits across diverse populations from 1000 genomes data using diverse imputation panels**
A) American admixed imputed versus the Haplotype Reference Consortium panel B) American admixed imputed versus the Consortium on Asthma among African-Ancestry Populations in the Americas (CAAPA) panel C) American admixed imputed versus the Genome Asia Pilot (GAsP) panel D) European imputed versus the Haplotype Reference Consortium panel E) African imputed versus the CAAPA panel F) African admixed imputed versus the CAAPA G) East Asians imputed versus the GAsP panel H) South Asians imputed versus the GAsP panel. Blue denotes Global Diversity Array + custom content imputation and red denotes Global Diversity Array imputation

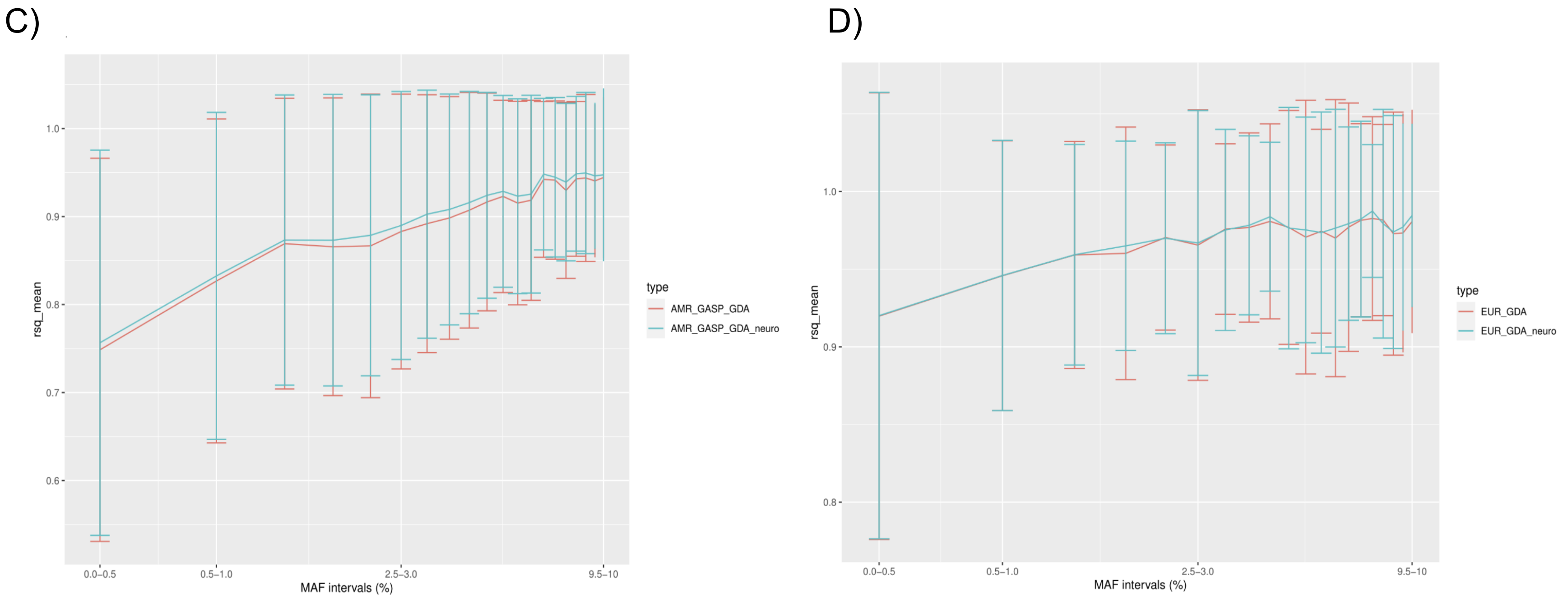


**Supplementary Figure 4A-H. Imputation accuracy of tag GWAS hits across diverse populations from 1000 genomes data using diverse imputation panels**
A) American admixed imputed versus the Haplotype Reference Consortium panel B) American admixed imputed versus the Consortium on Asthma among African-Ancestry Populations in the Americas (CAAPA) panel C) American admixed imputed versus the Genome Asia Pilot (GAsP) panel D) European imputed versus the Haplotype Reference Consortium panel E) African imputed versus the CAAPA panel F) African admixed imputed versus the CAAPA G) East Asians imputed versus the GAsP panel H) South Asians imputed versus the GAsP panel. Blue denotes Global Diversity Array + custom content imputation and red denotes Global Diversity Array imputation

E)

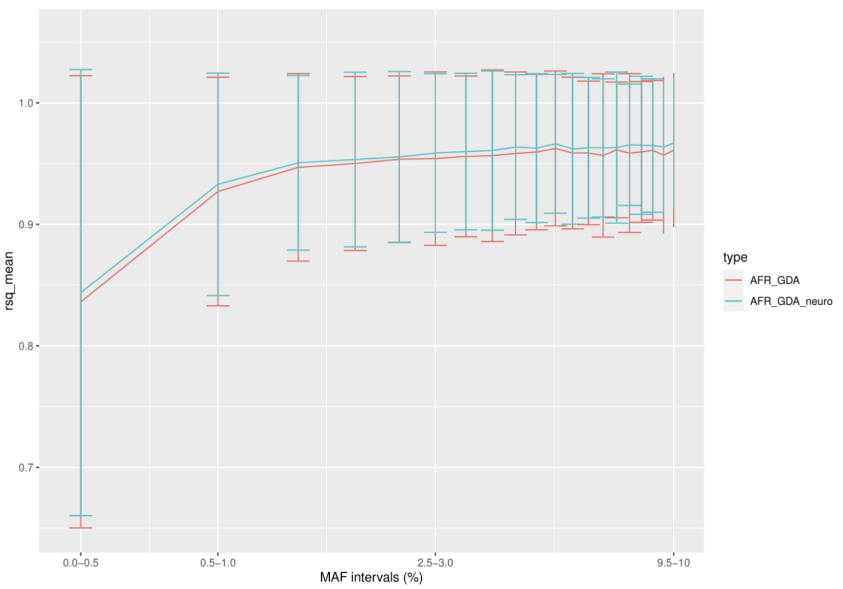


F)

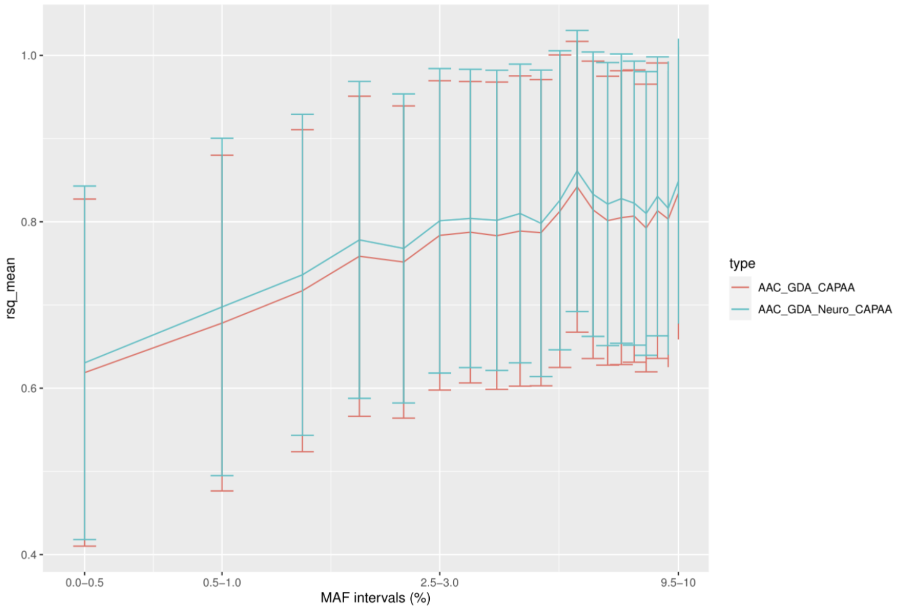


**Supplementary Figure 4A-H. Imputation accuracy of tag GWAS hits across diverse populations from 1000 genomes data using diverse imputation panels** A) American admixed imputed versus the Haplotype Reference Consortium panel B) American admixed imputed versus the Consortium on Asthma among African-Ancestry Populations in the Americas (CAAPA) panel C) American admixed imputed versus the Genome Asia Pilot (GAsP) panel D) European imputed versus the Haplotype Reference Consortium panel E) African imputed versus the CAAPA panel F) African admixed imputed versus the CAAPA G) East Asians imputed versus the GAsP panel H) South Asians imputed versus the GAsP panel. Blue denotes Global Diversity Array + custom content imputation and red denotes Global Diversity Array imputation

G)

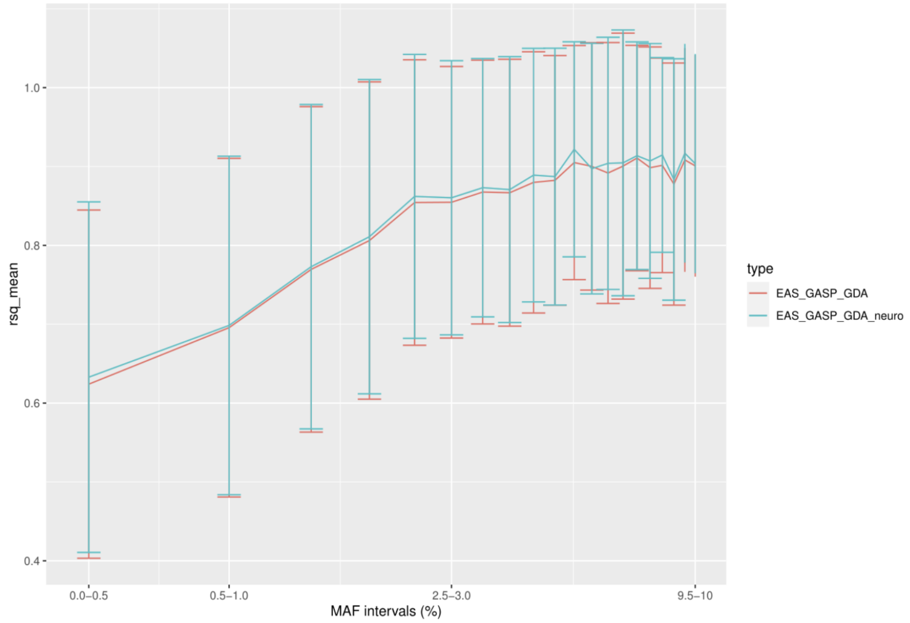


H)

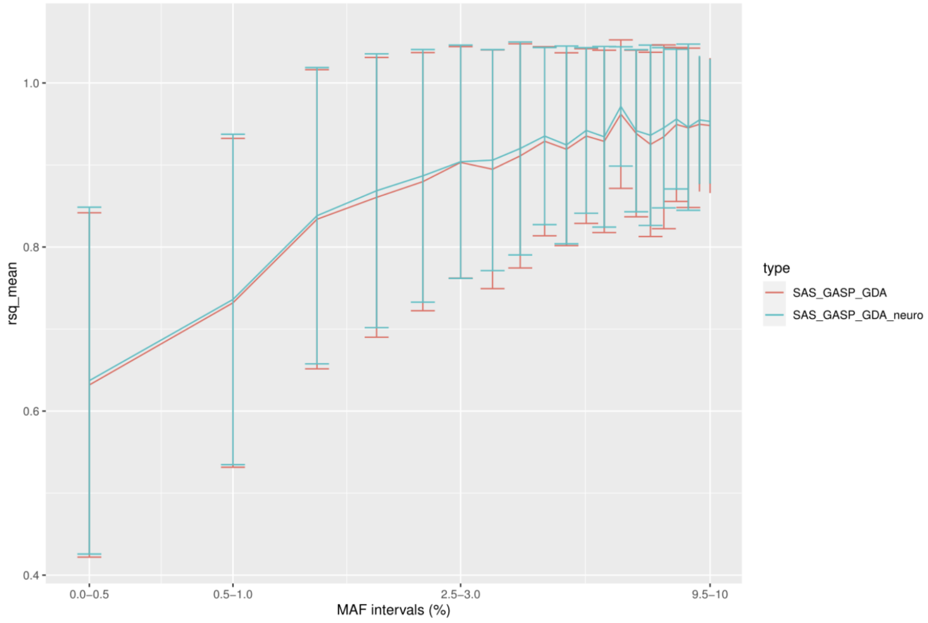


**Supplementary Figure 4A-H. Imputation accuracy of tag GWAS hits across diverse populations from 1000 genomes data using diverse imputation panels** A) American admixed imputed versus the Haplotype Reference Consortium panel B) American admixed imputed versus the Consortium on Asthma among African-Ancestry Populations in the Americas (CAAPA) panel C) American admixed imputed versus the Genome Asia Pilot (GAsP) panel D) European imputed versus the Haplotype Reference Consortium panel E) African imputed versus the CAAPA panel F) African admixed imputed versus the CAAPA G) East Asians imputed versus the GAsP panel H) South Asians imputed versus the GAsP panel. Blue denotes Global Diversity Array + custom content imputation and red denotes Global Diversity Array imputation
